# T cell tolerant fraction as a predictor of graft-vs-host disease following allogeneic hematopoietic cell transplantation

**DOI:** 10.1101/2025.11.08.25338304

**Authors:** Jared Ostmeyer, Andrew Cox, Elizabeth Nassar, Farrukh Awan, Amir Toor

## Abstract

**Background:** Donor T cell reconstitution following allogeneic hematopoietic cell transplantation (HCT) is crucial for protective immunity against infections and cancer relapse. However, its use remains limited by the risk of graft-versus-host disease (GvHD), a serious complication arising when donor T cells target recipient tissues. Here we evaluate a novel metric—the T cell receptor beta (TRB) tolerant fraction—as a predictor of GvHD risk.

**Methods:** We analyzed TRB sequencing data from mouse models of GvHD and pre-transplantation samples from previously published human HCT studies. We computed the TRB tolerant fraction based on the overlap between baseline *(pre-conditioning)* donor and recipient TRB repertoires and assessed its ability to predict acute and chronic GvHD.

**Results:** In mice, a common donor-recipient pairing (DRP) known to induce severe acute GvHD exhibited lower TRB tolerant fractions compared to control DRP with lower risk of GVHD. In human patients, individuals who developed acute GvHD showed a trend toward lower TRB tolerant fractions compared to those who remained free of GVHD. Notably, patients developing chronic GvHD had lower TRB tolerant fractions compared to patients without chronic GvHD (p = 0.019). ROC-AUC analysis further validated predictive capability for chronic GvHD (AUC = 0.81) in the cohort examined.

**Conclusions:** The TRB tolerant fraction calculated from pre-transplant donor-recipient TRB sequence overlap robustly predicts chronic GvHD and may do so for acute GvHD as well. This method may enhance donor selection and personalized risk assessment strategies, providing an avenue for reducing GvHD incidence through improved T cell compatibility screening and merits further study.

**Highlights:**

- We introduce the TRB tolerant fraction to measure of how many donor TCRβ sequences fall within a recipient’s tolerance space.
- The metric compares donor TRB sequences that overlap the recipient’s productive TRB repertoire *(i.e expressed and be tolerant selected)* with those overlapping the recipient’s non-productive TRB sequences *(cannot express and not tolerant selected)*.
- The metric is evaluated in established mouse donor–recipient pairs and in human allogeneic hematopoietic cell transplantation (allo-HCT) cohorts.
- We find the tolerant fraction shows a consistent signal associated with risk of graft-versus-host disease *(a clinical sign of failed T-cell tolerance)* in both mice and humans.
- The TRB tolerant fraction may add information beyond routine clinical factors.

## Introduction

Reconstitution of donor T cells following allogeneic hematopoietic cell transplantation (HCT) is integral to mediating beneficial immune responses, including the graft-versus-leukemia (GvL) effect, which reduces cancer relapse risk ^1^. Beyond antitumor responses, donor T cell recovery confers protection against severe viral infections, such as Epstein–Barr virus, adenovirus, and SARS-CoV-2 ^2–4^. Donor lymphocyte infusion (DLI), a procedure administering additional donor T cells post-transplantation, further highlights the therapeutic potential of donor T cells, achieving sustained remission in relapsed chronic myeloid leukemia (CML) and controlling residual disease across multiple hematologic malignancies ^5^.

A major obstacle restricting broader clinical implementation of HCT is the risk of severe alloreactivity, manifesting as graft-versus-host disease (GvHD). GvHD can occur when donor T cells recognize recipient tissues as foreign, leading to severe inflammation and damage in organs such as the skin, gastrointestinal tract, lungs and the liver. Minor histocompatibility antigen mismatches between HCT donors and recipients can induce strong, rapid, and often severe T cell mediated alloreactivity. Given the extensive variability in human leukocyte antigen (HLA) molecules across individuals, HCT without careful donor-recipient HLA matching can thus lead to severe alloreactivity or even fatal outcomes.

Current methods for reducing the risk of GvHD focus primarily on matching donor and recipient HLA types and pharmacologic immune suppression post-transplantation. However, HLA typing alone does not address the T cell repertoire of the donors, which may include T cell receptors (TCRs) with specificity for recipient antigens responsible for alloreactivity, potentially explaining why as many as 4 out of 10 HLA matched HCT can result in GvHD ^6^. Recent technological advancements have made large-scale sequencing of TCRs cost-effective and practical, enabling detailed analysis and comparison of donor and recipient T cell repertoires ^7^. By directly comparing these repertoires, it may be possible to assess donor-recipient compatibility at the TCR level, potentially enhancing prediction accuracy beyond traditional HLA-typing based approaches.

Previous research using TCR sequencing has identified associations between specific TCR patterns and the development of GvHD, yet these insights were derived from post-transplantation samples ^8–12^. To improve upon this approach, we propose a novel approach termed, the T cell receptor beta (TRB) tolerant fraction, recently utilized in predicting immune-related adverse events (irAE) ^13^. This method quantifies the compatibility of donor and recipient TCR repertoires, offering a novel pre-transplantation metric that may enhance the prediction of GvHD risk and inform clinical decisions in allo-HCT.

### T Cell Receptor Tolerant Fraction

We developed the TRB tolerant fraction to estimate how much of a donor’s TRB repertoire lies within a recipient’s self-tolerant space (Figure 1A). Starting from a recipient’s pre-transplant TRB repertoire, we partition sequences into *productive* (in-frame, no premature stops, and capable of expressing a functional TRB chain) and *non-productive* (out-of-frame or with premature stops) rearrangements. Productive TRB sequences from recipients are assumed to be self-tolerant because they can express a functional TRB chain that would have successfully undergone thymic selection. Non-productive TRB sequences, unable to produce functional proteins, would not have been selected for self-tolerance and therefore may be non-tolerant ^14–18^.

**Figure 1:**
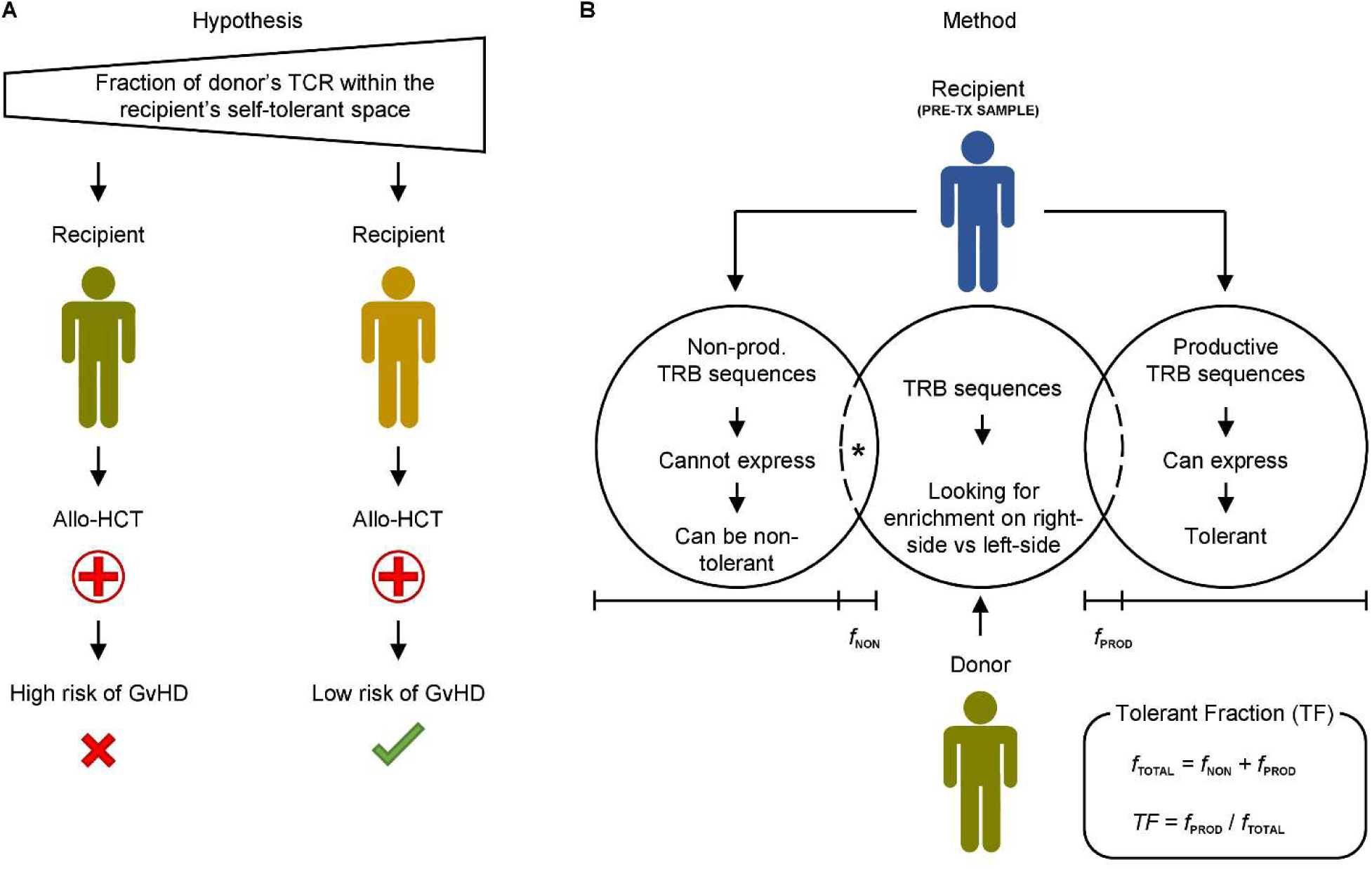
Hypothesis and methodology. (A) Hypothesis: In allogeneic HCT (allo-HCT), GvHD risk decreases as a larger share of donor TCRs lies within the recipient’s self-tolerant repertoire space, and increases when more donor TCRs fall outside it. (B) Method: The T cell receptor beta (TRB) tolerant fraction attempts to measure how much of the donor’s TRB repertoire lies within the recipient’s self-tolerant space. From a pre-transplant recipient sample, TRB sequences are split into two sets—non-productive *(cannot be expressed; potentially non-tolerant)* and productive *(expressible; presumed self-tolerant)*. Donor TRB sequences are compared with both sets to estimate how much of the donor repertoire aligns with the recipient’s tolerant space. The asterisk marks the in-silico repair of non-productive sequences to enable translation and like-for-like comparison with productive sequences *(see Methods)*.

To quantify overlap with the recipient’s tolerant space (Figure 1B), we compare the donor’s CDR3 (complimentary determining region 3) set, labelled *D*, with the recipient’s productive and non-productive CDR3 sets:

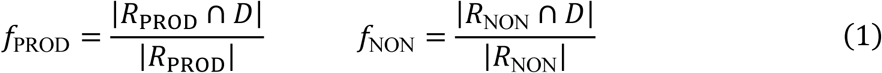

Here, *R*_PROD_ and *R*_NON_ are the sets of recipient’s unique CDR3 amino-acid sequences from productive and non-productive TRB rearrangements, *D* is the donor’s unique CDR3 set (productive or non-productive, as specified below), ∩ denotes exact, overlapping CDR3 matches, and ∣ *S* ∣ denotes cardinality of *S* (the number of unique sequences in *S*). We ignore template counts so that each sequence contributes once. In equations #1, the denominator is the size of the recipient set and the numerator is the portion that overlaps with the donor. Thus, *f*_PROD_ and *f*_NON_ are the proportions of the recipient’s *productive* (tolerant) and *non-productive* (potentially non-tolerant) repertoires shared with the donor.

To summarize whether overlap favors the tolerant space, we define the tolerant fraction:

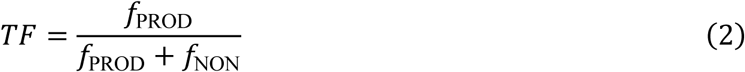

The tolerant fraction ranges from 0 to 1 *(e.g. 0% to 100%)*. Values near 1 indicate greater overlap with the recipient’s productive *(self-tolerant)* set. Values near 0 indicate greater overlap with the non-productive set. We hypothesize that higher tolerant fractions reflect lower GvHD risk because the donor lies within the recipient’s tolerant space to a greater extent. In this study, T cell repertoires from established mouse models of GvHD as well as human HCT were studied to assess the relationship between GvHD and the TRB tolerant fraction.

See Supplementary Figure 1 for the calculation overview and Supplementary Materials for an example TRB Tolerant Fraction.

## Methods and Materials

### Mouse Samples and TCR Sequencing

The TRB tolerant fraction was assessed for a well-characterized murine donor–recipient pairing (DRP) with established differences in GvHD outcomes, using existing splenic TRB repertoires rather than performing new transplantations ^19,20^. Specifically, we analyzed two DRP: C57BL/6→BALB/c (fully MHC-mismatched, severe GvHD expected) and the syngeneic control BALB/c→BALB/c (no GvHD expected). These pairings were selected to represent the extremes of alloreactivity—from severe to none—and to test whether the TRB tolerant fraction could distinguish between them. TRB repertoires were generated by sequencing unsorted splenocytes from BALB/c and C57BL/6 mice (Adaptive Biotechnologies, Seattle, WA ^21^). TRB sequencing data from six mice (four BALB/c and two C57BL/6) were obtained from publicly available datasets (https://doi.org/10.21417/ADPT2015MC and https://doi.org/10.21417/ADPT2019BTMC), as summarized in Table 1.

**Table 1:** Summary of mouse samples used for TRB sequencing analysis. Mouse strain identifiers, provider, age, sex, tissue sources *(spleen)*, and counts of unique productive *(expressed, tolerant)* and non-productive *(not expressed, potentially non-tolerant)* TRB sequences are shown.

| Unique TRB Gene Counts |  |  |  |  |  |
| --- | --- | --- | --- | --- | --- |
| Mouse | Provider | Age and Sex | Tissue | Productive | Non-productive |
| URL: https://doi.org/10.21417/ADPT2015MC |  |  |  |  |  |
| C57BL/6 #1 | Charles River Laboratories | 18 week-old males | Spleen | 49549 | 18533 |
| C57BL/6 #2 | Charles River Laboratories | 18 week-old males | Spleen | 49964 | 18593 |
| BALB/c #1 | Charles River Laboratories | 18 week-old males | Spleen | 59477 | 18347 |
| BALB/c #2 | Charles River Laboratories | 18 week-old males | Spleen | 58882 | 18614 |
| URL: https://doi.org/10.21417/ADPT2019BTMC |  |  |  |  |  |
| BALB/c #3 | Jackson Laboratories | 10–11 week-old females | Spleen | 17029 | 6546 |
| BALB/c #4 | Jackson Laboratories | 10–11 week-old females | Spleen | 21480 | 7890 |

### Human Samples, and TCR Sequencing, and Clinical Outcomes

We also assessed the clinical relationship between the pre-transplantation TRB tolerant fraction and GvHD outcomes using two published studies with paired donor and recipient TRB repertoires. The first study was an interventional study using post-transplantation cyclophosphamide and was approved by the Johns Hopkins Medicine IRB and/or the FHCRC/University of Washington Cancer Consortium IRB ^22^. The second study was prospective and non-interventional, approved by Cleveland Clinic IRBs #5024 and #4927 with written informed consent ^8^. We included sequences from only the first HCT with both donor and recipient pre-transplantation TRB repertoires and no T cell depletion. We used only samples collected ≥28 days before conditioning. Donors were either haploidentical (n=6) or HLA-matched related donors (n=13). Patient ages ranged from 25-66 years, and donor ages ranged from 32 to 67. Conditioning was either myeloablative (MAC, n=15), or reduced intensity (RIC, n=4). Conditioning regiments used were: Busulfan/Cyclophosphamide (n=7), Busulfan/Fludarabine (n=6), Fludarabine/Cyclophosphamide/total body irradiation (n=2), or Fludarabine/total body irradiation (n=4). Underlying diseases included: AML (n=12), BAL (n=2), MDS/AML (n=1), MF (n=2), and CML (n=2). Prophylaxis included: Post Transplant Cyclophosphamide (n=4), cyclosporine/mycophenolate mofetil (n=5), tacrolimus/mycophenolate/Cyclophosphamide (n=5), FK/MMF/MTX (n=4), FK/Cy (n=1). Patients were followed for at least 365 days or until death. Acute GVHD (n=8) was graded 0–IV and chronic GVHD (n=8) was captured retrospectively from chart review.

### Calculating the TRB tolerant fraction

All TRB sequencing was performed by Adaptive Biotechnologies (Seattle, WA) ^21^. Details of how TRB sequences are represented and translated for these overlaps is as follows. Because TCR function is determined by protein sequence, all TRB rearrangements were translated to predicted amino-acid sequences. Productive sequences translate directly. For non-productive sequences, we applied a published in-silico “repair” algorithm that makes minimal edits *(delete 1–2 nucleotides at junctions or substitute a nucleotide)* to restore the reading frame or remove premature stops, generating one or more plausible protein sequences per rearrangement ^15^. When multiple repairs were possible, we retained all plausible repaired sequences for analysis. Full algorithmic details are provided elsewhere ^13^, which is strictly computational. As these repaired sequences bypass thymic selection, they are expected to be enriched for specificities that would be removed in vivo (recipient-intolerant). All calculations in Eq. (1) used only the CDR3 region of each TRB protein sequence.

Finally, we selected the donor set *D* in Eq. (1) to match the GvHD context. For acute GvHD *(mouse models and human allo-HCT),* productive TRB sequences were used for *D* in equations #1, which may represent donor T cells actually transferred to the recipient and capable of driving acute GvHD. For chronic GvHD studies, donor non-productive sequences were used for *D*, which have not undergone thymic selection in the donor and may reflect newly generated donor-derived T cells arising from donor hematopoietic stem cells (HSCs) after transplant. This choice is motivated by mouse studies suggesting chronic GvHD can occur when donor-derived T cells develop in the recipient without prior donor thymic selection ^23, 24^. We acknowledge this is a simplification but consider it a practical starting point.

### Statistical Analysis

Statistical analyses tested specific hypotheses related to the TRB tolerant fraction. For mouse models, our null hypothesis stated that the TRB tolerant fraction in non-GvHD (C57BL/6→C57BL/6, BALB/c→BALB/c) and mild-GVHD DRP (BALB/c→C57BL/6) was not higher than the TRB TF in severe GvHD pairings (C57BL/6→BALB/c). Similarly, in human studies, the null hypotheses stated that the TRB tolerant fractions in patients without acute GvHD (grades 0–1) or without chronic GvHD was not higher than in patients with severe acute GvHD (grades 2–4) or chronic GvHD. The alternative hypothesis for each comparison was that the TRB tolerant fraction was higher in groups without severe GvHD. Since only this directional hypothesis was tested, we employed a one-sided Mann-Whitney U test.

## Results

### Mouse Models

We first evaluated the TRB tolerant fraction in mouse models to determine whether our method could accurately identify a known donor-recipient pairing (DRP) with severe acute GvHD. The C57BL/6 (H-2^b^) → BALB/c (H-2^d^) transplant model, a fully MHC-mismatched pairing with robust acute GvHD, was used as our positive control (figure 2A) ^19, 20,25^. Using spleen-derived TRB sequences from these mice, we calculated a TRB tolerant fraction for 8 possible DRP (Equations #1 & 2), resulting in a mean value of 58.2% *(maximum 59.4%)* (figure 2B). For controls, we assessed genetically matched syngeneic histo-compatible *BALB/c → BALB/c* DRP, representing ideal conditions with minimal GvHD risk. The syngeneic models exhibited tolerant fractions higher than the severe GvHD setting, resulting in a mean value of 61.1% (*minimum 59.8%)*. These findings are consistent with the hypothesis that a lower TRB tolerant fraction corresponds to greater alloreactivity and higher GvHD risk.

**Figure 2:**
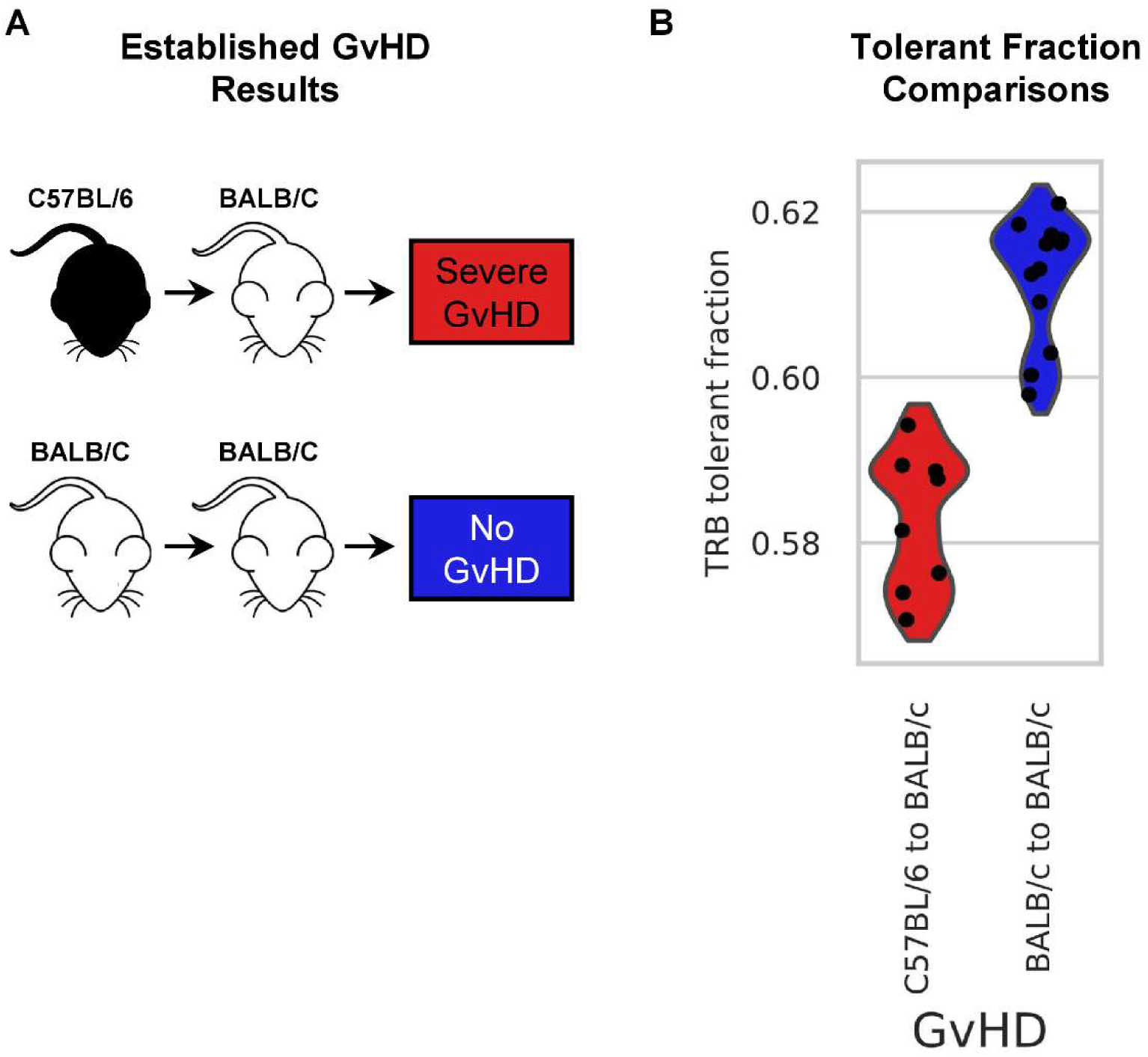
Relationship between TRB tolerant fraction and GvHD severity in mouse models. (A) Schematic overview of murine transplantation models: The fully MHC-mismatched C57BL/6 → BALB/c model reliably induces severe acute GvHD (red), whereas control pairings (BALB/c → BALB/c) show no GvHD. (B) Violin plots of the TRB tolerant fraction for each donor-recipient pairing show significantly lower tolerant fractions in the severe GvHD model compared to control pairings (blue), consistent with the hypothesis.

*Results for the DRP C57BL/6 → C57BL/6 and BALB/c → C57BL/6 are provided in Supplementary Figure 2*.

### Acute GvHD in Human HCT recipients

We next analyzed TRB sequences from previously published human studies that provided pre-transplantation TRB sequences from DRP, alongside documented acute GvHD outcomes (Table 2). Nineteen HCT recipients were included, of whom 11 remained free of severe acute GvHD (grades 0–1), while 8 developed acute GvHD (grades 2– 4). Using equations #1-2 with the pretransplant donor and recipient TRB repertoires, we calculated the TRB tolerant fractions for each DRP. Patients who did not develop severe acute GvHD had higher tolerant fractions than patients who did not (mean 85.2%, std 5.0%) compared to those who did (mean 82.6%, std 5.7%), although this difference did not reach statistical significance at the current sample size (p = 0.087) (figure 3A). By applying a threshold to the TRB tolerant fraction, we attempted to distinguish between the two outcomes *(free of acute GvHD vs acute GvHD)*, allowing us to generate a receiver operating characteristics (ROC) curve. The area under the curve (AUC) curve was 0.69, with the true positive rate consistently above the false positive rate (figure 3B).

**Figure 3:**
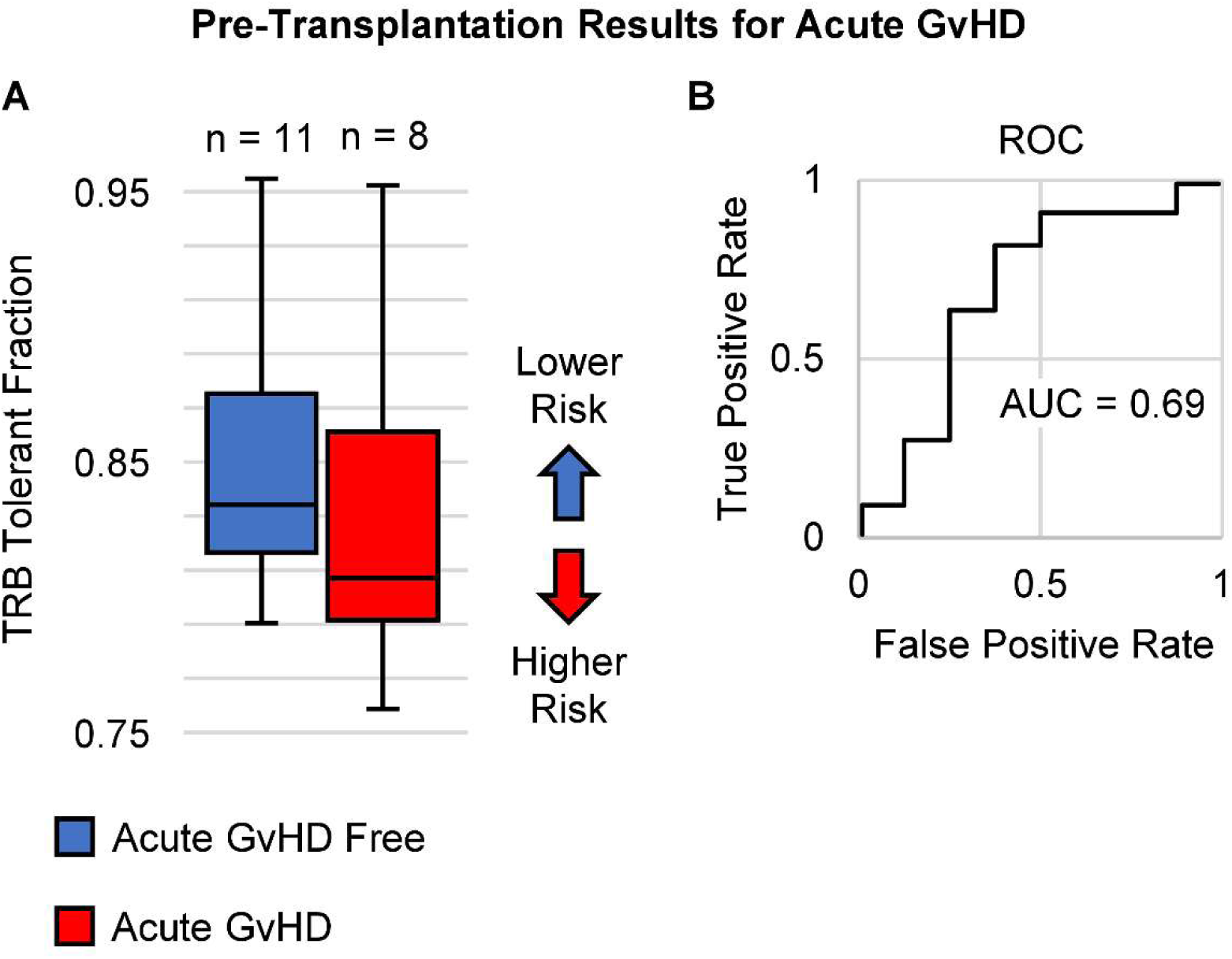
Pre-transplantation TRB tolerant fraction as a predictor of acute GvHD in human patients. (A) Box plot comparing TRB tolerant fractions between patients who remained free of acute GvHD (blue, n = 11) and patients who developed acute GvHD (red, n = 8). Patients without acute GvHD generally exhibited higher tolerant fractions, indicating lower predicted risk. (B) Receiver operating characteristic (ROC) curve demonstrating the performance of the TRB tolerant fraction in predicting acute GvHD. The area under the curve (AUC) is 0.69, indicating moderate predictive ability.

**Table 2:** Clinical characteristics of patients undergoing an allo-HCT, including disease type, donor type, acute and chronic GvHD outcomes, and counts of unique productive (Prod.) and non-productive (Non.) TRB sequences collected for both donors and recipients. Disease abbreviations: AML (acute myeloid leukemia), BAL (biphenotypic acute leukemia), MDS (myelodysplastic syndrome), MF (myelofibrosis), CML (chronic myeloid leukemia). Donor type abbreviations: Haplo (haploidentical donor), MRD (matched related donor). Follow-up durations marked with an asterisk (*) indicate patients not observed long enough for definitive chronic GvHD outcomes.

| Study | Disease | Donor Type | Patient Age | Donor Age | Conditioning regimen | GVHD prophylaxis | Intensity Conditioning | GvHD |  | # Unique TCRβs Collected |  |  |  |
| --- | --- | --- | --- | --- | --- | --- | --- | --- | --- | --- | --- | --- | --- |
|  |  |  |  |  |  |  |  | Acute | Chronic | Donor |  | Recipient |  |
|  |  |  |  |  |  |  |  |  |  | Prod. | Non. | Prod. | Non. |
| JCI Insight 2016 | AML | MRD | 40s | 30s | Bu/Flu | PTCy | MAC | N | N | 49348 | 9888 | 42909 | 8387 |
|  | AML | MRD | 60s | 60s | Bu/Flu | PTCy | MAC | Y | N | 39796 | 9187 | 31855 | 6789 |
|  | AML | MRD | 30s | 40s | Bu/Flu | PTCy | MAC | N | N | 16769 | 3929 | 66368 | 13883 |
|  | AML | MRD | 30s | 30s | Bu/Flu | PTCy | MAC | Y | N | 67309 | 15092 | 3679 | 787 |
| JCI Insight 2021 | AML | MRD | 50s | 40s | Bu/Cy | FK/MMF/MTX | MAC | Y | N | 6340 | 1444 | 23954 | 4000 |
|  | BAL | Haplo | 60s | 40s | Flu/Cy/TBI-200cGy | FK/MMF/Cy | RIC | Y | Y | 46370 | 8557 | 63409 | 10915 |
|  | MDS/AML | Haplo | 60s | 30s | Flu/TBI | FK/Cy | MAC | N | Censor | 2297 | 468 | 9476 | 1702 |
|  | AML | MRD | 50s | 50s | BuCy | CSA/MMF | MAC | Y | N | 2827 | 592 | 7626 | 1902 |
|  | AML | MRD | 40s | 50s | Bu/Cy | FK/MMF/MTX | MAC | N | N | 15646 | 3059 | 37252 | 6977 |
|  | MF | MRD | 50s | 60s | Bu/Cy | FK/MMF/MTX | MAC | N | Y | 25157 | 4548 | 918 | 159 |
|  | MF | MRD | 50s | 50s | Bu/Cy | FK/MMF/MTX | MAC | N | Y | 91495 | 15653 | 6847 | 1054 |
|  | BAL | Haplo | 20s | 30s | Flu/TBI | FK/MMF/Cy | MAC | N | N | 53096 | 9632 | 27023 | 5066 |
|  | AML | MRD | 40s | 50s | Bu/Flu | CSA/MMF | RIC | Y | Y | 16011 | 2921 | 14227 | 2829 |
|  | CML | Haplo | 30s | 40s | Flu/TBI | FK/MMF/Cy | MAC | N | Y | 2907 | 605 | 37717 | 7398 |
|  | AML | MRD | 60s | 50s | Bu/Flu | CSA/MMF | RIC | Y | Y | 21185 | 3857 | 52526 | 10268 |
|  | AML | Haplo | 60s | 30s | Flu/Cy/TBI | FK/MMF/Cy | RIC | N | Censor | 18210 | 3255 | 16482 | 3032 |
|  | AML | Haplo | 30s | 60s | Flu/TBI | FK/MMF/Cy | MAC | N | N | 21167 | 3956 | 8925 | 1842 |
|  | CML | MRD | 50s | 50s | Bu/Cy | CSA/MMF | MAC | N | Y | 36666 | 5725 | 4205 | 750 |
|  | AML | MRD | 40s | 40s | Bu/Cy | CSA/MMF | MAC | Y | Y | 20896 | 4407 | 821 | 192 |

### Chronic GvHD in Human HCT recipients

Finally, we examined chronic GvHD using the same cohort and pre-transplantation TRB sequences, now limited to 17 patients with documented chronic GvHD outcomes (Table 2). Among these patients, 9 remained free of chronic GvHD while 8 developed chronic GvHD. Once again, using equations #1 & 2 with the pre-transplant donor and recipient TRB repertoires, we obtained TRB tolerant fractions for each DRP *(This calculation differs from the acute analysis, which used donor productive rather than non-productive sequences, as described earlier)*. The tolerant fraction was significantly higher in patients who did not develop chronic GvHD (mean 59.6%, std 2.1%) compared to those who did (mean 51.3%, std 2.0%; p = 0.019, Mann-Whitney U test) (figure 4A). Applying a tolerant fraction threshold to classify chronic GvHD outcomes, we found that most potential cut-off values resulted in a true positive rate exceeding the false positive rate. Specifically, using a threshold of 68.7%, 7 out of 9 patients without chronic GvHD were correctly classified (78% sensitivity), and all 8 patients with chronic GvHD fell below this threshold (100% specificity). The overall predictive performance, as quantified by receiver operating characteristic (ROC) analysis, yielded an area under the curve (AUC) of 0.81 (figure 4B).

**Figure 4:**
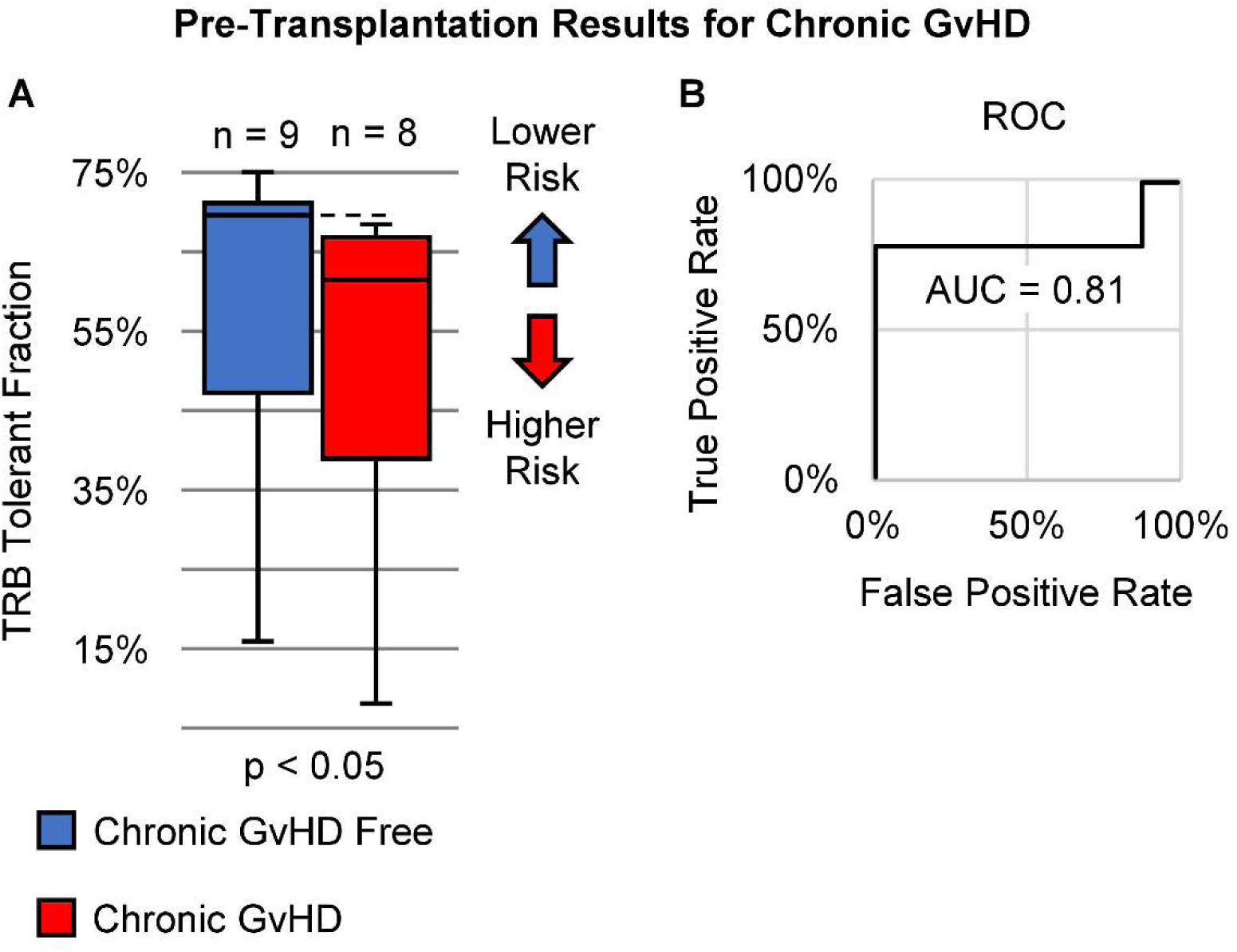
Pre-transplantation TRB tolerant fraction effectively predicts chronic GvHD in human patients. (A) Box plot illustrating significantly higher TRB tolerant fractions in patients who remained free of chronic GvHD (blue, n = 9) compared to patients who developed chronic GvHD (red, n = 8), indicating lower risk with higher tolerant fractions (p < 0.05, Mann-Whitney U test). **(B)** Receiver operating characteristic (ROC) curve demonstrates strong predictive capability for chronic GvHD, with an area under the curve (AUC) of 0.81.

## Discussion

Despite advances in transplantation protocols, around 40% of recipients still develop GvHD ^6^, a complication that can affect nearly any organ, often with severe symptoms. In some cases, GvHD can be fatal. Beyond its clinical severity, GvHD also imposes significant emotional and financial burdens, with median healthcare costs for managing severe chronic GvHD ranging between approximately $280,000 and $657,000 ^26, 27^. Although recent advances like post-transplant cyclophosphamide and abatacept have reduced GvHD incidence ^28–30^, this complication continues to impact clinical outcomes, emphasizing the urgent need for better predictive and preventive measures.

In response to this ongoing clinical challenge, our study investigates the potential of using TRB sequencing to predict GvHD risk. Previous efforts to profile GvHD have largely focused on post-transplant measures such as T cell diversity, clonality, and similarity, none of which effectively predict GvHD from pre-transplant samples. Moreover, these metrics do not directly reflect the biological interactions underlying T cell alloreactivity and GvHD, limiting their predictive utility. To overcome these limitations, we propose the TRB tolerant fraction as a potential predictive measure for GvHD. In our approach, the TRB tolerant fraction attempts to computationally assesses hypothetical donor T cell compatibility with the recipient prior to transplantation, potentially identifying patients at heightened risk of GvHD. Ultimately, our goal is to enable safer and broader use of HCT, both for cancer and other therapeutic contexts.

In the C57BL/6→BALB/c mouse model, the TRB tolerant fraction differentiated severe GvHD from syngeneic controls, supporting the biological basis of the approach. In pre-transplant human samples, higher tolerant fractions were observed with lower rates of both acute and chronic GvHD. While the relationship with acute GvHD did not achieve statistical significance *(likely due to the small sample size and low event rate)*, our analysis considered only the TRB tolerant fraction, without accounting for clinical factors such as age, sex, disease type, CMV status, conditioning, and transplantation protocols, which are known to strongly influence acute GvHD. Larger cohorts and multivariable analyses are needed to account for these variables. By contrast, the chronic-GvHD results were statistically significant, highlighting what may be achievable with a more comprehensive approach. Most patients who remained free of the disease showed notably higher tolerant fractions than those who developed chronic GvHD. Notably, a few outliers with unusually low tolerant fractions had limited genomic material for sequencing, potentially explaining their anomalous results.

Of note is the relatively modest separation (3–8%) between tolerant fraction values both in the mouse model and in patients who developed GvHD compared to those who remained free of the disease. Interestingly, a similarly narrow range was observed when applying the TRB tolerant fraction to predict immune-related adverse events (irAEs) in our previous study, as well as in other studies suggesting only a few hundred clones expand significantly during these adverse events ^31^. This observation could suggest that only a small fraction of donor T cells contribute significantly to GvHD risk. This is consistent with the Power Law distribution of T cell clonal frequencies observed in previous studies ^32^. This distribution is characterized by a small number of high frequency clones, accompanied by a much larger number of lower frequency T cell clones, a pattern replicated by antigen binding affinity driven T cell clonal growth models ^33^. Therefore given the multitude of T cell clones identified in each normal repertoire, it is plausible that the numerically small differences observed result in clinically meaningful alloreactivity.

Importantly, the TRB tolerant fraction is not intended to replace traditional HLA typing in donor selection. Instead, the TRB tolerant fraction complements HLA typing, as each method addresses one aspect of the TCR-mHA-HLA interaction. Larger studies integrating HLA typing and the TRB tolerant fraction through comprehensive computational and algorithmic approaches will be essential to fully validate this combined predictive strategy.

In conclusion, our study identifies the TRB tolerant fraction as associated with future chronic GvHD risk, and with potential predictive value for acute GvHD as well. Importantly, these predictions are based entirely on pre-transplantation data, underscoring the potential clinical utility of this method during donor selection and prophylactic planning. Additionally, the TRB tolerant fraction may help clinicians identify patients needing closer post-transplant monitoring. In this scenario, the TRB tolerant fraction could complement, not replace HLA typing, as each method addresses one side of the TCR-HLA interaction. We believe that integrating the TRB tolerant fraction into clinical practice could significantly enhance the safety and applicability of allo-HCT, thereby facilitating broader use of donor T cells for treating cancer and other diseases.

## Data Availability

All data produced in the present study are available upon reasonable request to the authors

## Abbreviations

allo-HCT: allogeneic hematopoietic cell transplantation
allo-HSCT: allogeneic hematopoietic stem cell transplantation
AML: acute myeloid leukemia
AUC: area under the ROC curve
BAL: biphenotypic acute leukemia
BALB/c: inbred mouse strain used in transplant models
Bu: busulfan (conditioning)
Bu/Flu: busulfan + fludarabine (conditioning regimen)
Bu/Cy: busulfan + cyclophosphamide (conditioning regimen)
C57BL/6: inbred mouse strain used in transplant models
CDR3: complementarity-determining region 3 of the TCRβ chain
CML: chronic myeloid leukemia
CMV: cytomegalovirus
Cy: cyclophosphamide (conditioning/prophylaxis)
D: donor set of unique CDR3 sequences (equation variable)
DLI: donor lymphocyte infusion
DRP: donor–recipient pair (pairing)
fNON: fraction of R_NON overlapping with D
fPROD: fraction of R_PROD overlapping with D
FHCRC: Fred Hutchinson Cancer Research Center
FK: tacrolimus (FK-506)
Flu: fludarabine (conditioning)
GvHD: graft-versus-host disease
GvL: graft-versus-leukemia
H-2^b: murine MHC haplotype of C57BL/6
H-2^d: murine MHC haplotype of BALB/c
Haplo: haploidentical donor
HCT: hematopoietic cell transplantation
HLA: human leukocyte antigen
HSC: hematopoietic stem cell
IRB: Institutional Review Board
irAE: immune-related adverse event
MAC: myeloablative conditioning
MDS: myelodysplastic syndrome
MF: myelofibrosis
MHC: major histocompatibility complex
MMF: mycophenolate mofetil
MRD: matched related donor
MTX: methotrexate
Non.: non-productive TRB sequences (table label)
PTCy: post-transplant cyclophosphamide
Prod.: productive TRB sequences (table label)
R_NON: recipient set of unique non-productive CDR3 sequences
R_PROD: recipient set of unique productive CDR3 sequences
RIC: reduced-intensity conditioning
ROC: receiver operating characteristic
SARS-CoV-2: severe acute respiratory syndrome coronavirus 2
Syngeneic: genetically identical donor–recipient pairing (e.g., BALB/c→BALB/c)
TBI: total body irradiation
TCR: T-cell receptor
TF: tolerant fraction
TRB: T-cell receptor beta chain
|S|: cardinality of set S (number of distinct sequences)
∩: set intersection (intersection of two sets)
Flu/Cy/TBI: fludarabine + cyclophosphamide + total body irradiation (conditioning regimen)
Flu/TBI: fludarabine + total body irradiation (conditioning regimen)
FK/MMF/MTX: tacrolimus + mycophenolate mofetil + methotrexate (GVHD prophylaxis)
FK/Cy: tacrolimus + cyclophosphamide (GVHD prophylaxis)

## Consent to Participate

This study analyzed only de-identified, publicly available human TCRβ sequencing data from previously published cohorts. No new human subjects were recruited, contacted, or intervened upon. The original human studies were conducted under Institutional Review Board (IRB) approved by either the Johns Hopkins Medicine IRB, the FHCRC/University of Washington Cancer Consortium IRB or the Cleveland Clinic IRBs and with informed consent, as described in the method section ^8, 22^.

## Animal Research Statement

No new animal experiments were performed for this manuscript. We analyzed already published mouse TRB repertoires.

