## Supplementary Materials for "T cell tolerant fraction as a predictor of graft-vs-host disease following allogeneic hematopoietic cell transplantation"

### Supplemental Figures

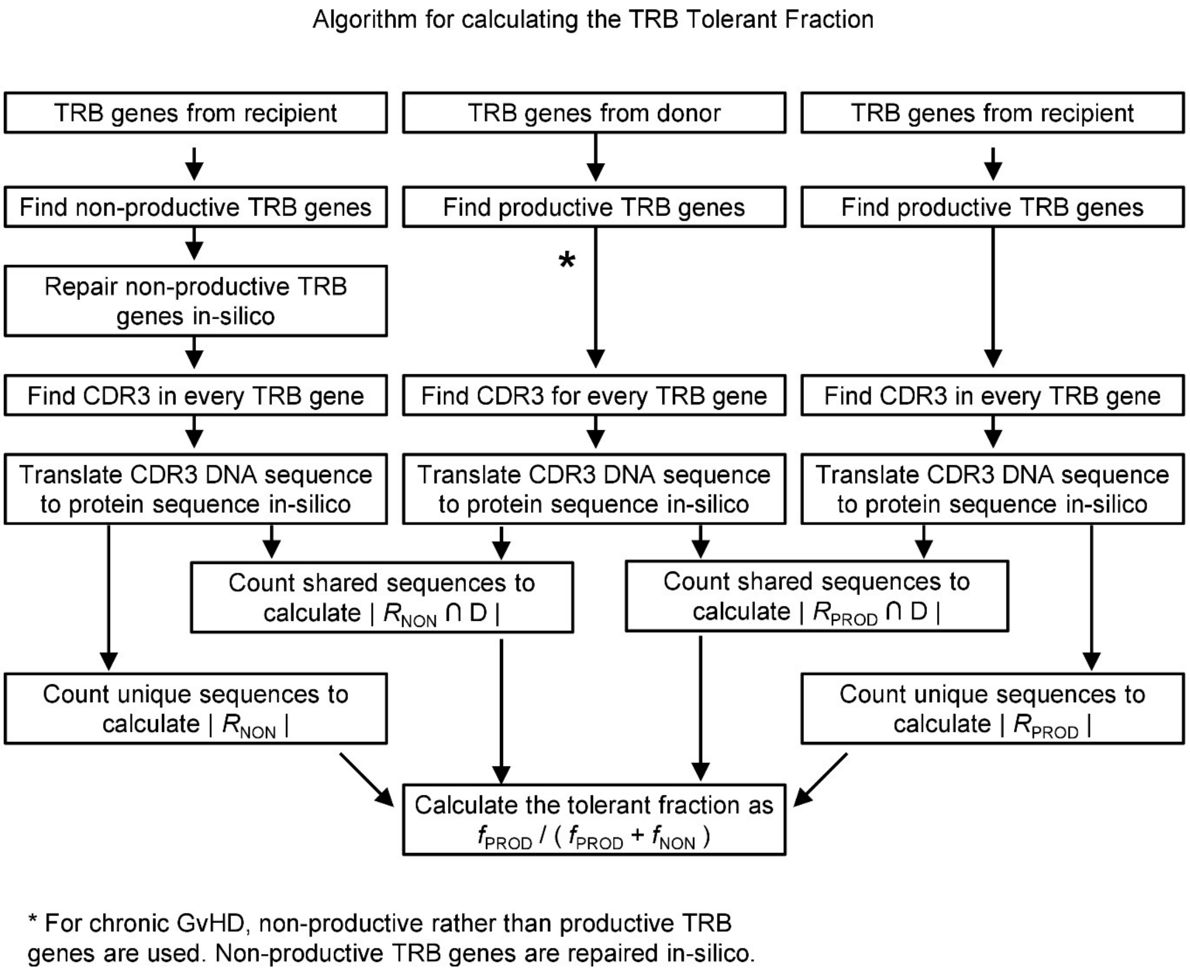


**Supplemental Figure 1:** Flow chat diagram of the algorithm for calculating the TRB tolerant fraction.

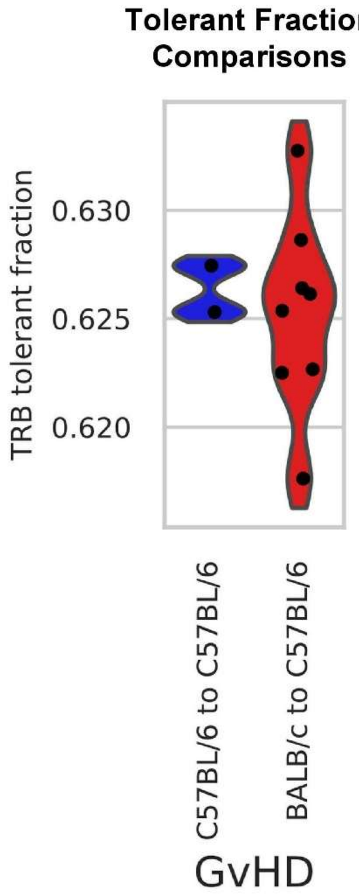


**Supplemental Figure 2:** Violin plots of the TRB tolerant fraction for each donor-recipient pairing of our dataset mice for C57BL/6 → C57BL/6 and BALB/c → C57BL/6.

### Supplemental Text

#### Example Calculation of TRB Tolerant Fraction

To illustrate how the TRB Tolerant Fraction is calculated, a toy example using a very number of small donor and recipient repertoires is presented below. In practice, the same procedure is run at scale on full repertoires. We begin with the CDR3 nucleotide sequences and their productive or non-productive status.

*Donor Repertoire:*

1. TGTGCCAGCAGAGGTGAAGTTGACACC Prod
2. TGTGCCAGCAGTTTCAGACGGTCTGGCTACACCTTC Prod
3. TGTGCCACCGACGGCGGGAAATAC Prod

*Recipient Repertoire:*

a. TGTGCCAGCAGTCTGCAGTACTTC Prod
b. TGTGCCAGCAGGGGGGAGGTTGACACC Prod
c. TGTGCCAGCAGTGACGGGACTTAGTAC Non-prod
d. TGTGCCACCGTATGGAGGGAAGTAC Non-prod

Next, we translate to the DNA nucleotide sequences to amino acid sequences. Productive sequences translate codon-by-codon to amino acid sequences. Non-productive sequences are "repaired" in silico with minimal edits *(delete 1–2 nucleotides at junctions or substitute a single nucleotide)* to restore the reading frame or remove premature stops, yielding one or more plausible amino-acid (AA) sequences per rearrangement. Because the junctional residues are stochastically generated, it is assumed that any edits could have occurred naturally, justifying these minimal edits. Junctional residues can be identified by searching for nucleotides not matching germline encoded sequences, and in this example are indicated by an "*"

*Productive Donor Repertoire:*

1. TGTGCCAGCAGAGGTGAAGTTGACACC — Prod
   C A S R G E V D T

2. TGTGCCAGCAGTTTCAGACGGTCTGGCTACACCTTC — Prod
   C A S S F R R S G Y T F

3. TGTGCCACCGACGGCGGGAAATAC — Prod
   C A T D G G K Y

*Productive Recipient Repertoire:*

a. TGTGCCAGCAGTCTGCAGTACTTC — Prod
   C A S S L Q Y F

b. TGTGCCAGCAGGGGGGAGGTTGACACC — Prod
   C A S R G E V D T

c. TGTGCCAGCAGTGACGGGACTTAGTAC — Non-Prod *(Stop codon TAG)*
   *** * — Junctional

TGTGCCAGCAGTGACGGGACTAAGTAC — Mutate T to A
C A S S D G T K Y

TGTGCCAGCAGTGACGGGACTCAGTAC — Mutate T to A
C A S S D G T Q Y

TGTGCCAGCAGTGACGGGACTGAGTAC — Mutate T to A
C A S S D G T E Y

d. TGTGCCACCGTATGGAGGGAAGTAC — Non-Prod *(out-of-frame)*
   **** * — Junctional

TGTGCCACCXTATGGAGGGAAGTAC — Delete G *(in-frame)*
C A T Y G G K Y

TGTGCCACCGXATGGAGGGAAGTAC — Delete T *(in-frame)*
C A T D G G K Y

TGTGCCACCGTXTGGAGGGAAGTAC — Delete A *(in-frame)*
C A T V G G K Y

TGTGCCACCGTAXGGAGGGAAGTAC — Delete T *(in-frame)*
C A T V G G K Y

TGTGCCACCGTATGGAGGXGAGTAC — Delete G *(in-frame)*

C A T V W R E Y

Summarizing the amino acid sequences, we have:

*Donor Repertoire:*

1. CASRGEVDT Prod
2. CASSFRRSGYTF Prod
3. CATDGGKY Prod

*Recipient Repertoire:*

a. CASSLQYF Prod

b. CASRGEVDT Prod

c. CASSDGTKY, CASSDGTQY, CASSDGTQY Non-prod

d. CATYGGKY, CATDGGKY, CATVGGKY, CATVGGKY, CATVWREY Non-prod

Multiple valid repairs can inflate the number of non-productive derived amino acid sequences, so using fractions of shared sequences $f_{\text{PROD}}$ and $f_{\text{NON}}$ from Eq. (1) keeps the contributions balanced. As defined in the manuscript, let $R_{\text{PROD}}$ be the set of unique recipient sequences from productive rearrangements, $R_{\text{NON}}$ be the set of unique recipient sequences derived from non-productive rearrangements after minimal repairs, and $D$ be the donor set of unique sequences *(productive or non-productive depending on the clinical context)*. For out toy example, we have:

| | |
|---|---|
| $\lvert R_{\text{PROD}} \rvert = 2$ | (e.g. CASSLQYF and CASRGEVDT) |
| $\lvert R_{\text{PROD}} \cap D \rvert = 1$ | (e.g. CASRGEVDT in both donor and recipient) |
| $\lvert R_{\text{NON}} \rvert = 6$ | (e.g. CASSDGTKY, CASSDGTQY, CATYGGKY, CATDGGKY, CATVGGKY, and CATVWREY; igoring duplicates) |
| $\lvert R_{\text{NON}} \cap D \rvert = 1$ | (e.g. CATDGGKY) |

With these values, we calculate $f_{\text{PROD}}$ and $f_{\text{NON}}$ as:

$$f_{\text{PROD}} = \frac{|R_{\text{PROD}} \cap D|}{|R_{\text{PROD}}|} = \frac{1}{2} = 0.5$$

$$f_{\text{NON}} = \frac{|R_{\text{NON}} \cap D|}{|R_{\text{NON}}|} = \frac{1}{6} = 0.17$$

Finally, we calculate the TBR Tolerant Fraction as:

$$TF = \frac{f_{\text{PROD}}}{f_{\text{PROD}} + f_{\text{NON}}} = \frac{0.5}{0.5+0.17} \approx 0.75 = 75\%$$
